# Automated Identification of Patients with Immune-related Adverse Events from Clinical Notes using Word embedding and Machine Learning

**DOI:** 10.1101/2020.05.19.20106583

**Authors:** Samir Gupta, Anas Belouali, Neil J Shah, Michael B Atkins, Subha Madhavan

## Abstract

Immune Checkpoint Inhibitors (ICIs) have substantially improved survival in patients with advanced malignancies. However, ICIs are associated with a unique spectrum of side effects termed Immune-Related Adverse Events (irAEs). To ensure treatment safety, research efforts are needed to comprehensively detect and understand irAEs from real world data (RWD). The goal of this work is to evaluate a Machine Learning-based phenotyping approach that can identify patients with irAEs from a large volume of retrospective clinical notes representing RWD. Evaluation shows promising results with an average F1-score=0.75 and AUC-ROC=0.78. While the extraction of any available irAEs in charts achieves high accuracy, individual irAEs extraction has room for further improvement.

## Introduction and Background

Immune checkpoint inhibitors (ICIs) have substantially transformed outcomes for patients with many advanced malignancies [1]. FDA-approved ICIs include ipilimumab, nivolumab, pembrolizumab, atezolizumab, avelumab, durvalumab and cemiplimab [2]. These therapies boost the immune system to kill tumor cells by blocking immunosuppressive molecules expressed on immune cells such as cytotoxic T-lymphocyte antigen 4 (CTLA-4), and programmed cell death 1 (PD-1), or other cells within the tumor microenvironment such as programmed cell death ligand 1 (PD-L1). However, by increasing the activity of the immune system, ICIs can induce a range of inflammatory side effects termed Immune-related Adverse Events (irAEs) [2,3].

The incidence of irAEs of any grade in patients treated with nivolumab, pembrolizumab, atezolizumab, durvalumab, avelumab and cemiplimab is 26.8% [4]. irAEs occur in a high proportion (up to 61%) of patients treated with ipilimumab, a drug that is increasingly used in combination with nivolumab in the treatment of a variety of cancers [5]. Although any organ system can be affected, irAEs most commonly involve the gastrointestinal tract, skin, endocrine glands, and liver [2,4]. irAEs are pathophysiologically different from the toxicities of conventional cancer drugs and because they involve activation of physiologic processes the predisposing factors are less well defined [2]. Complications from ICIs can potentially be severe and may diminish the benefits in outcomes from using ICIs [4]. Implementation of irAE management algorithms may help patients to obtain benefit from CPI therapy while minimizing the impact of irAEs [6]. Hence, research efforts are needed to identify and comprehensively characterize irAEs in order to provide improved AE management for patients receiving ICIs [7].

With the widespread adoption of Electronic Health Records (EHRs), a significant amount of clinical data is recorded about patients, including their diagnosis, medications, and encounters. Such electronic databases are being promoted as “real world evidence” to study efficacy and safety of new and existing treatments [8–10]. However, knowledge about adverse events in general and irAEs specifically is mostly buried in clinical narratives in these electronic databases. Also, manually extracting information from patients’ charts is time-consuming, costly, and lacks standardization [11]. To ensure patient safety, it is important to adequately record these irAEs in EHR and related clinical systems, such as sponsor databases for clinical trials. Methodologies that could automate the recording of irAEs from EHRs would be beneficial to accurately annotate datasets for secondary use of EHR data such as for adverse event registries and for other research purposes. Additionally, it would benefit accurate cohort identification for clinical trials and accelerate the creation of curated datasets for developing predictive models. In this study, we specifically focus on clinical notes of cancer patients treated with ICIs. Our goal is to automate the identification of patients who developed any type of irAE and detect the most common irAE subtypes and their grades. We hypothesize that an automated approach using word embeddings and Machine Learning (ML) on a set of longitudinal clinical notes of patients treated with ICIs could rapidly identify patients who developed irAEs and detect their individual irAEs. Furthermore, extraction and standardization of irAEs from EHRs can serve as a gold standard to augment efforts to standardize irAEs from other sources such as FDA drug labels[12]. Hsiehchen et al. [13] have shown that there is discordance among how clinicians diagnose and characterize irAEs from cancer immunotherapy.

Various tools and methodologies have been developed to extract patient information from unstructured clinical text using Natural Language Processing (NLP) and Machine Learning. Clinical NLP pipelines such as CLAMP [14], cTakes [15], and CliNER [16] have been developed to extract named-entities such as problems, diagnosis, treatments, labs, and medications from clinical charts. Researchers have also employed supervised and semi-supervised ML models for classifying text in clinical notes with different clinical phenotypes. Li et al. [17] used a deep learning model for extracting adverse drug events from clinical notes and achieved an F1-score of 65.9%. Gehrmann et al. [18] compared concept extraction based methods with Convolutional neural networks (CNN) and other common classification models in ten different phenotyping tasks (e.g. advanced cancer, substance abuse, chronic pain, obesity, etc.) using discharge summaries. They showed that CNNs outperform concept extraction-based methods in almost all of the phenotyping tasks, with an improvement in F1-score of up to 26%. Banerjee et al. [19] proposed a weakly supervised NLP pipeline to classify sentences in clinical notes with patient-centered outcomes following prostate cancer treatment. In this work, sentence vectors based on a weighted function of word embeddings were used as input to logistic models (F1-score = 0.86). Wang et al. [20] used weakly labeled data automatically generated from a rule-based algorithm with various ML models for classifying text snippets in clinical notes with smoking status and proximal femur (hip) fracture labels. Other works in the area of classification of text in notes include: detecting heart failure [21], silent brain infarcts from neuroimaging reports [22], bleeding disorders/events [23] and ureteric stones in radiology reports [24].

In this work, we describe a supervised ML-based NLP pipeline to classify patients treated with ICIs as having irAEs or not based on their longitudinal set of clinical notes. We specifically compare the performance of different ML models (shallow and deep learning models) using different feature representations (frequency-based, distributed word embeddings) and text reduction (keyword-based filtering) methods. Previous works have focused on note-level or sentence-level classification for detecting clinical phenotypes. To the best of our knowledge, this is the first study to use all clinical notes to identify irAEs at the patient-level. Our work demonstrates that deep learning models with text-reduction using keyword filtering and word embeddings can achieve good accuracy in classifying patients with irAEs based on the clinical narratives.

## Materials and Methods

We present an overall workflow of the study design and methodologies in Fig 1.

**Fig 1.**
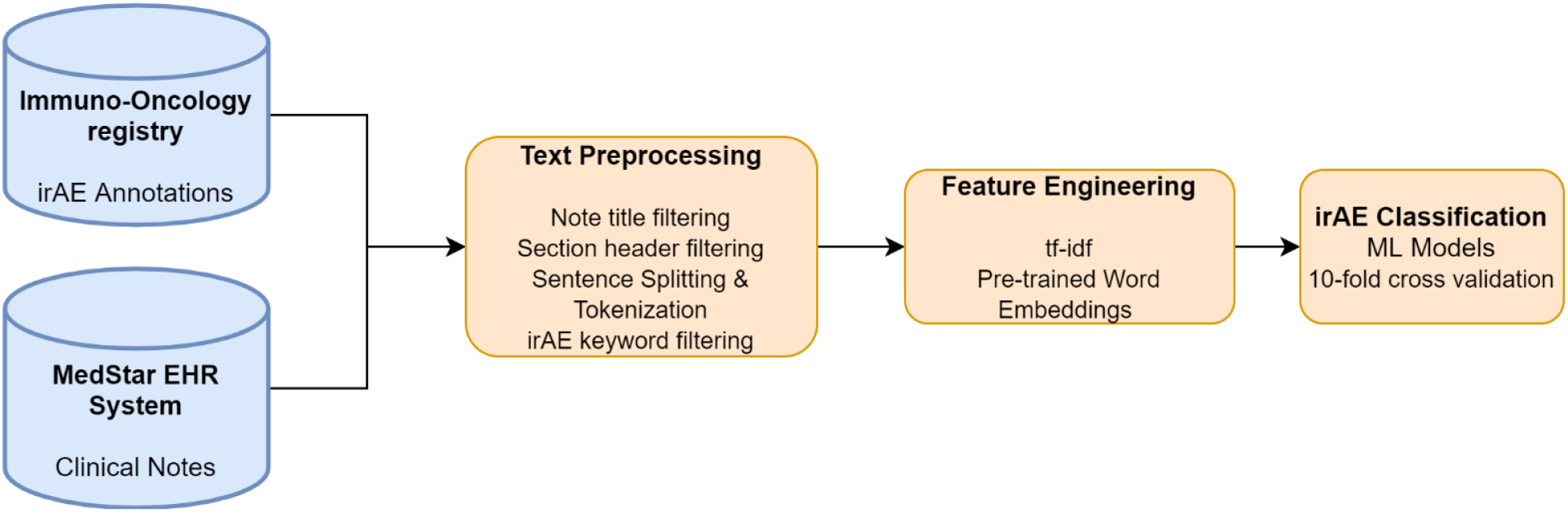
System Workflow. Abbreviations used: tf-idf (term frequency-inverse document frequency)

### Clinical Setting and Data Source

We used the EHR system of MedStar Health as the data source for the study. MedStar Health is a non-profit healthcare organization of over 120 entities including 10 hospitals in the Baltimore-Washington metropolitan area, which provides care for more than four million people each year. Our team of clinicians and informaticians built a centralized research repository for Immuno--Oncology designed for investigators with HIPAA compliant access to enable novel hypothesis generation and retrospective outcomes research. The registry includes demographics, diagnoses, laboratories, medication, treatment administration and toxicity events for about 758 patients treated with different ICIs from January 2011 to April 2018 at 5 Hospitals; MedStar Georgetown University Hospital (MGUH), MedStar Washington Hospital Center (MWHC), Franklin Square Hospital (FSH), Good Samaritan Hospital (GSH), and Union Memorial Hospital (UMH). Our team of clinicians extracted irAEs (immunotherapy toxicity) from the EHRs by conducting a retrospective and prospective chart review of patients with advanced cancer treated with ICI agents. Since patients can be treated with multiple non-overlapping ICI therapies, we have 818 patient-ICI therapy pairs in the Immuno-Oncology registry. Fig 2 depicts the breakdown of toxicities in the curated Immuno-Oncology registry (tx, pts indicates patient-therapy pair and patients respectively). The incidence and severity of irAEs were identified per the National Cancer Institute’s Common Terminology Criteria for Adverse Events (CTCAE) v4.03 guidelines [25]. The mean age of the cohort is 66.51 (std=13.43). The gender breakdown of the cohort is 58.4% Male, 41.6% Female. The race breakdown of the patients is: White (469, 61.9%), Black or African American (202, 26.6%), Asian (21, 2.8%), American Indian or Alaska Native (1, 0.1%), Not Specified/Unknown (32, 4.2%), Other (33, 4.4%).

**Fig 2.**
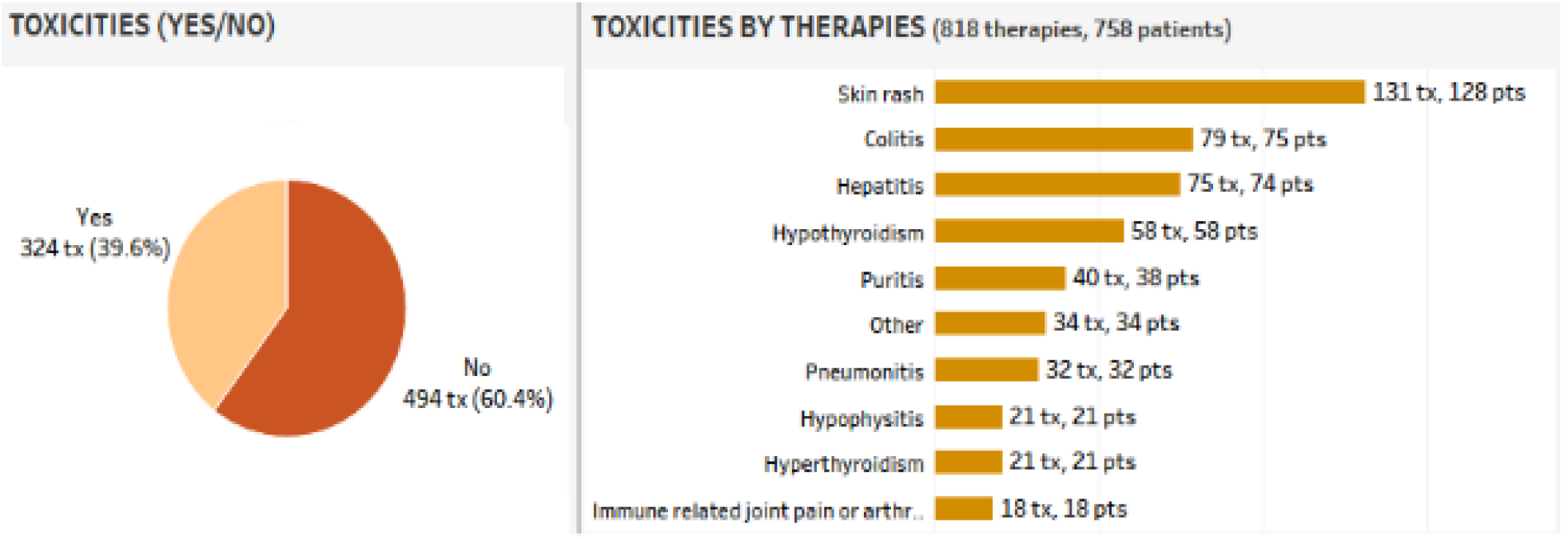
irAEs toxicity breakdown in the Immuno-Oncology registry. Abbreviations used: tx: patient-therapy pairs; pts: patients

### Data Collection and Preparation

From the Immuno-Oncology registry, we identified 724 patients treated with 781 therapies that had at least one clinical text note during their ICI treatment duration. We selected all clinical notes between the date of the first dose of the ICI and the last date of the follow-up or date of start of another ICI. Selecting notes beyond the date of the last dose will allow us to account for delayed irAEs that can happen several weeks or even months after the date of the last dose [26]. 9924 notes were then extracted from the Oncology EHR at MedStar and de-identified using the De-ID software [27] to remove all patient health identifiers. Every ICI therapy for a patient was reviewed by clinicians and annotated for irAEs. irAEs occurred in 43.8 % of patients (42.9 % of therapies) in the database. We decided to merge certain related irAEs subtypes in order to have a considerable number of patients/therapies with positive irAEs. “Skin rash”, “pruritus” and “other skin toxicities” annotations were combined under the individual irAE subtype “Skin-related toxicity”, while “Hypothyroidism”, “Hyperthyroidism”, “Hypophysitis” and “other endocrine toxicities” annotations were combined under irAE subtype “Endocrine toxicity”. The top 3 irAEs in our dataset for which we had at least one clinical notes are (1) skin-related toxicity (skin rash, pruritus, etc.) in 20.3 % patients (19.3 % of therapies), (2) Endocrine toxicities in 12.8 % of patients (11.2 % of therapies) and (3) Colitis in 10.4 % of patients (10 % of therapies). We will refer to each of the 781 patient-ICI therapy pairs as patients henceforth for simplicity.

### Text Preprocessing

As indicated in the above section, for each patient, we extracted clinical notes between the date of the first dose of the ICI and the last date of the follow-up or the start date of another ICI. Our task was to classify each patient based on their set of notes, as having developed any type of irAEs or not. We also built 3 additional classifiers to identify patients with the top three irAEs (skin-related toxicity, colitis, endocrine toxicity). To reduce the number of clinical notes associated with each patient, we performed certain preprocessing steps such as filtering based on note titles, section headers, and the presence of irAE keywords.

The evidence of irAEs as stated in clinical notes is typically present in certain note types and sections. We developed a tool to extract title and section headers from a clinical note. The first non-empty line of a note is extracted as the note title. We used simple regular expressions to extract section headers and text associated with each section. All note titles and section headers were extracted from the set of 9924 notes and provided to a clinician for feedback. We asked the clinician to indicate title and section headers that are most likely to contain statements of the patient developing an irAE. Examples of relevant note titles include: “Medical Oncology Note”, “Progress Note”, “Hematology Progress Note” etc. and relevant section headers include: “History of Present Illness (HPI)”, “Assessment and Plan”, “Review of Systems” etc. Based on the set of title and header filters, we reduced the number of clinical notes and associated text for further processing. An example sentence of irAE evidence (“colitis”) in the HPI section of a clinical note is provided below in Example 1.

#### Example 1

*“developed diarrhea ….and I prescribed prednisone … daily for probable ipilimumab induced diarrhea”*

We used NLTK [28] to split the text into sentences and tokenize each sentence into words. We noticed that certain information in clinical notes for a patient was replicated across notes. This is due to the fact that physicians typically modify the previous visit note for a patient when creating a new note. To alleviate this duplicate information issue, we kept only unique sentences across all notes for a patient. Even though the amount of text was reduced by around 30% on average for a patient, we noticed a considerable amount of “noise” (sentences without irAE evidence). For example, sentences such as in Examples 2 and 3 are very common in Medical Oncology Notes. We hypothesize that removing such “noise” would improve performance of the Machine Learning models both in terms of accuracy and training time.

#### Example 2

*“Pt. he initially presented w/ 2.2 cm l shoulder mole and underwent shave bx …”*

#### Example 3

*“the pt was subsequently enrolled in clinical trial 701 (denileukin) and completed 4 cycles”*

In a recent work [12], the authors built a standardized database of irAEs that could serve as a gold standard for automatic irAE extraction. The database contains 893 irAE terms extracted from FDA drug labels for FDA-approved ICIs. We also asked a clinician to provide us with irAE terms for the top three irAEs in our Immuno-Oncology registry. We combined these sets of irAE keywords and selected only sentences that had a mention of an irAE. Note, irAE keyword-based filtering does not eliminate the need of an ML model. A keyword-only based approach will also select sentences which contain (1) irAEs that might be due to some other reason (not due to the ICI), (2) negative finding of irAE (e.g.: “patient denies rash”), or (3) a caution text (e.g.: “patient was advised of ipi related toxicities such as colitis..”). An ML model is still needed to discriminate between such negative mentions of irAEs and positive ones. Furthermore, to test our hypothesis of noise-removal based on irAE keywords, we report results with and without keyword filtering. A full list of irAE keywords along with the list of relevant titles and headers can be found at: https://github.com/samirgupta/irAE-classfication/blob/master/supplementary_file.xlsx. Finally, we combine all the selected sentences from the notes for a given patient in a longitudinal manner to yield a single text for each patient.

### Feature Engineering

To apply machine learning models, the text associated with clinical notes for a patient needs to be converted to structured features. The first set of features, which we used as input for our shallow ML models is term frequency-inverse document frequency (tf-idf) weighting [29]. The second set of features, which we used as input to our deep learning models are distributed word representations called word embeddings, which have been shown to achieve better performance in NLP tasks by learning similar vectors for similar words [30,31]. For our deep learning models, we used the publicly available pre-trained word embeddings from NCBI: BioWordVec [32,33]. BioWordVec are biomedical word embeddings, trained on PubMed and clinical notes from the MIMIC-III Clinical Database [34] using fastText [35]. The word vectors have the dimensionality of 200. Words which were not present in the set of pre-trained words are set as a zero vector.

### Classification Models

We employed several shallow ML and deep learning models to classify a patient treated with ICIs having developed any irAE and also the top three irAEs (skin-related toxicity, colitis, endocrine toxicity). The different shallow ML models with tf-idf feature representation that we tried are Logistic Regression (LR), Support Vector Machine (SVM), and Random Forest (RF). Additionally, we used pre-trained word embeddings (BioWordVec) to build and test two deep learning architectures, namely, Convolutional Neural Network (CNN) and Bi-directional Long Short-Term Memory (bi-LSTM).

The CNN architecture had a one-dimensional convolutional layer with rectified linear unit (ReLU) activation. For the convolutional layer, we experimented with three windows sizes: 3, 5 and 7, each of which has 400 filters. Every filter performs convolution on the text matrix and generates variable-length feature maps. We got the best results with a single window of size 5. Global max pooling was then performed over each map, which essentially extracts fixed-length global features for the text. A dense layer of size 1 with a sigmoid activation function was applied over the global features to obtain the CNN classifier. The biLSTM architecture consisted of a 64-cell bidirectional LSTM layer followed by two pooling layers (maximum and average). The maximum and average pooling layers were concatenated fed to a fully connected layer with 64 units (with ReLU activation). A dense layer of size 1 with a sigmoid activation function was applied over the fully connected layer to obtain the biLSTM classifier. For the deep learning models, we used binary cross-entropy as the objective loss function and the Adam algorithm [36] to optimize the loss function. To train the parameters for CNN and bi-LSTM, we used mini-batch training with a batch size of 32. In between each layer of our deep learning architectures, we added a dropout layer with a dropping probability of 0.5 to avoid overfitting during training. We set the number of epochs to 30 during training. For the classical ML models default parameters and loss function were used.

### Textual Analysis

We further analyzed the text using Scattertext [37], a text visualization tool to understand differences in clinical notes between patients having developed irAEs and those who didn’t. The tool uses a scaled f-score, which takes into account the category-specific precision and term frequency. While a term may appear frequently in both groups, the scaled f-score determines if the term is more characteristic of one category versus another. We excluded stopwords (such as “the”, “a”, “an”, “in”) from the text corpus.

## Results

### Toxicity Classification ML Results

We evaluated the performance of the binary classifiers to predict four toxicities labels: any irAEs (io-toxicity), and the top three irAEs (skin-related toxicity, colitis, and endocrine toxicity) using precision, recall, F1-score, and area under the receiver operating characteristic Curve (AUC-ROC) as performance metrics. Table 1 shows the class distribution (positive vs. negative instances) of the four toxicities.

**Table 1.** Class distribution of the toxicities.

| **Toxicity Label** | **Positive instances** | **Negative Instances** |
| --- | --- | --- |
| **Any irAE (io-toxicity)** | 335 (42.9%) | 446 (57.1%) |
| **Skin-related toxicity** | 152 (19.5%) | 629 (80.5%) |
| **Endocrine toxicity** | 93 (11.9%) | 688 (88.1%) |
| **Colitis** | 79 (10.1%) | 702 (89.9%) |

We used 10-fold cross-validation when computing the evaluation metrics. For each toxicity, all classifiers were evaluated with and without irAE keyword filtering (KF). For the shallow ML models, tf-idf was used as the features, while pre-trained word vectors (WV) from BioWordVec [32,33] were used as features for the deep learning models. Additionally, since the class distributions for the individual irAEs are imbalanced, we used the “class-weight” (CW) parameter to weight the cross-entropy loss function of the deep learning models to account for the class imbalance. 10-fold cross-validation results for any io-toxicity, skin-related toxicity, endocrine toxicity, and colitis are shown in Tables 2, 3a, 3b, and 3c respectively. Row 1 of the Tables 2 and 3 contains evaluation metrics for detecting irAEs with keyword filtering only, without applying any ML modelling. This will serve as our baseline mimicking a naive keyword search approach, which tags patients having irAEs, if any of the clinical notes associated with the patient contain an irAE mention. Note, KF for the individual irAEs (skin, endocrine, colitis) considers only irAE keywords corresponding to the individual irAEs.

**Table 2.** Evaluation results for irAEs (any io-toxicity)

| **Model** | **Approach** | **Precision** | **Recall** | **F1-score** | **AUC-ROC** |
| --- | --- | --- | --- | --- | --- |
| **None (baseline)** | KF | 42.9 | 100 | 60.0 | 50.0 |
| **LR** | tf-idf | 83.2 | 54.9 | 65.9 | 73.2 |
|  | KF+tf-idf | 81.1 | 57.3 | 66.9 | 73.4 |
| **SVM** | tf-idf | 77.3 | 60.1 | 67.9 | 73.4 |
|  | KF+tf-idf | 76.8 | 64.1 | 69.7 | 74.4 |
| **RF** | tf-idf | 80.0 | 59.1 | 67.8 | 73.4 |
|  | KF+tf-idf | 79.8 | 57.3 | 66.4 | 72.9 |
| **CNN** | WV | **84.9** | 63.5 | 71.2 | 77.3 |
|  | WV+KF | 81.5 | **69.8** | **75.2** | **78.2** |
| **biLSTM** | WV | 77.8 | 67.1 | 71.5 | 76.4 |
|  | WV+KF | 76.0 | 69.5 | 71.9 | 75.8 |

**Table 3a.** Evaluation results for skin-related toxicity.

| **Model** | **Approach** | **Precision** | **Recall** | **F1-score** | **AUC-ROC** |
| --- | --- | --- | --- | --- | --- |
| **None (baseline)** | KF | 28.5 | 98.2 | 44.1 | 69.3 |
| **LR** | tf-idf | 71.7 | 11.8 | 20.0 | 55.4 |
|  | KF+tf-idf | **78.4** | 46.7 | 57.3 | 71.6 |
| **SVM** | tf-idf | 70.2 | 34.2 | 45.4 | 65.3 |
|  | KF+tf-idf | 65.2 | 53.8 | 58.1 | 73.3 |
| **RF** | tf-idf | 70.0 | 10.5 | 18.1 | 54.8 |
|  | KF+tf-idf | 72.7 | 40.2 | 51.4 | 68.2 |
| **CNN** | WV | 76.5 | 44.0 | 53.2 | 70.1 |
|  | WV+KF | 73.6 | 59.8 | **65.1** | 77.1 |
|  | WV+KF+CW | 66.4 | **63.0** | 64.4 | **77.5** |
| **biLSTM** | WV | 68.6 | 46.1 | 55.1 | 70.5 |
|  | WV+KF | 74.1 | 53.2 | 60.2 | 74.3 |
|  | WV+KF+CW | 65.2 | 61.3 | 61.9 | 76.4 |

**Table 3b.** Evaluation results for endocrine toxicity.

| **Model** | **Approach** | **Precision** | **Recall** | **F1-score** | **AUC-ROC** |
| --- | --- | --- | --- | --- | --- |
| **None (baseline)** | KF | 20.3 | 100.0 | 33.8 | 73.5 |
| **LR** | tf-idf | 0 | 0 | 0 | 50.0 |
|  | KF+tf-idf | **85.5** | 20.5 | 31.7 | 59.9 |
| **SVM** | tf-idf | 67.5 | 18.1 | 27.1 | 58.7 |
|  | KF+tf-idf | 69.8 | 45.9 | 54.4 | 71.5 |
| **RF** | tf-idf | 0 | 0 | 0 | 50.0 |
|  | KF+tf-idf | 81.7 | 19.3 | 30.8 | 59.5 |
| **CNN** | WV | 83.0 | 25.6 | 36.7 | 62.6 |
|  | WV+KF | 73.4 | 44.2 | 53.2 | 70.8 |
|  | WV+KF+CW | 64.8 | 58.1 | **59.3** | 76.4 |
| **biLSTM** | WV | 74.2 | 33.2 | 43.0 | 65.7 |
|  | WV+KF | 65.8 | 54.7 | 58.4 | 75.0 |
|  | WV+KF+CW | 58.1 | **62.4** | 58.8 | **77.7** |

**Table 3c.** Evaluation results for colitis.

| **Model** | **Approach** | **Precision** | **Recall** | **F1-score** | **AUC-ROC** |
| --- | --- | --- | --- | --- | --- |
| **None (baseline)** | KF | 12.7 | 100.0 | 22.5 | 61.2 |
| **LR** | tf-idf | 50.0 | 7.8 | 13.3 | 53.9 |
|  | KF+tf-idf | 97.5 | 34.4 | 48.5 | 67.2 |
| **SVM** | tf-idf | 100.0 | 38.0 | 54.1 | 69.0 |
|  | KF+tf-idf | 84.1 | 55.7 | 65.8 | 77.2 |
| **RF** | tf-idf | 50.0 | 7.5 | 12.9 | 53.7 |
|  | KF+tf-idf | **95.8** | 36.9 | 50.6 | 68.3 |
| **CNN** | WV | 76.7 | 31.9 | 43.6 | 65.9 |
|  | WV+KF | 87.3 | 54.5 | 65.6 | 76.7 |
|  | WV+KF+CW | 78.6 | **67.1** | **70.9** | **82.3** |
| **biLSTM** | WV | 88.0 | 37.1 | 52.0 | 68.1 |
|  | WV+KF | 89.6 | 52.1 | 63.7 | 75.6 |
|  | WV+KF+CW | 73.2 | 63.4 | 65.2 | 79.8 |

For classifying irAEs (any io-toxicity), the naive rule-based keyword search approach (KF; row 1 of Table 1) results in 100% recall but precision of only 42.9%. This indicates that there was at least one clinical note for a patient with irAE that contained an irAE keyword, but only 42.9% of patients classified as having irAEs (at least one clinical note with an irAE keyword) had actually developed toxicity. Deep learning models yielded the best performance (F1-score and AUC-ROC) for classifying irAEs relative to shallow ML models. ML models on text reduced with keyword filtering tend to decrease precision slightly and increase recall with an overall increase in F1-score. Fig 3a and 3b depicts the f1 and AUC-ROC scores across the different models. CNN with KF yielded the best results (F1-score: 75.2% and AUC-ROC: 78.2%) for classifying irAE (any io-toxicity).

**Fig 3a.**
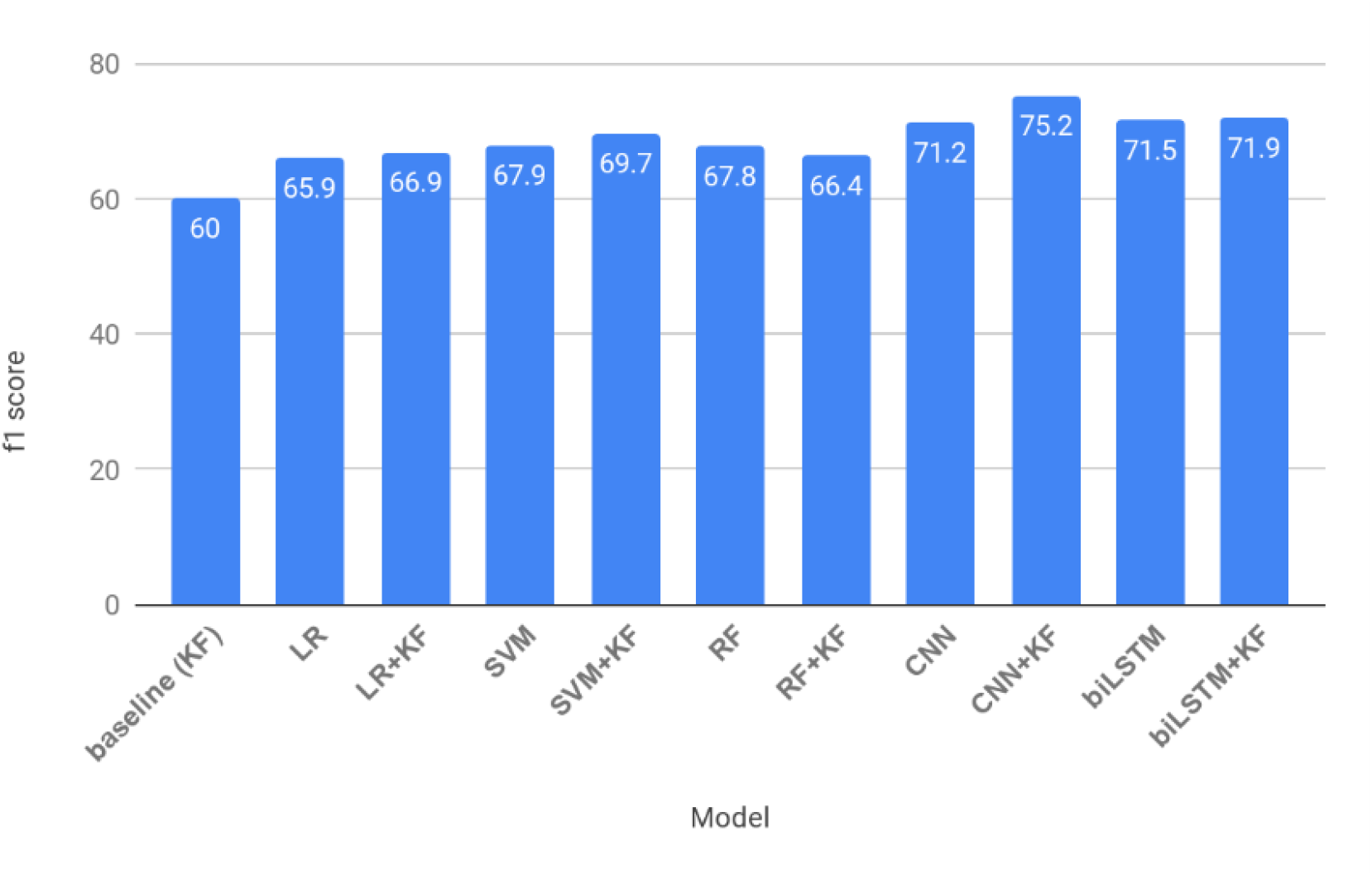
F1-score vs. ML models for classifying irAEs.

**Fig 3b.**
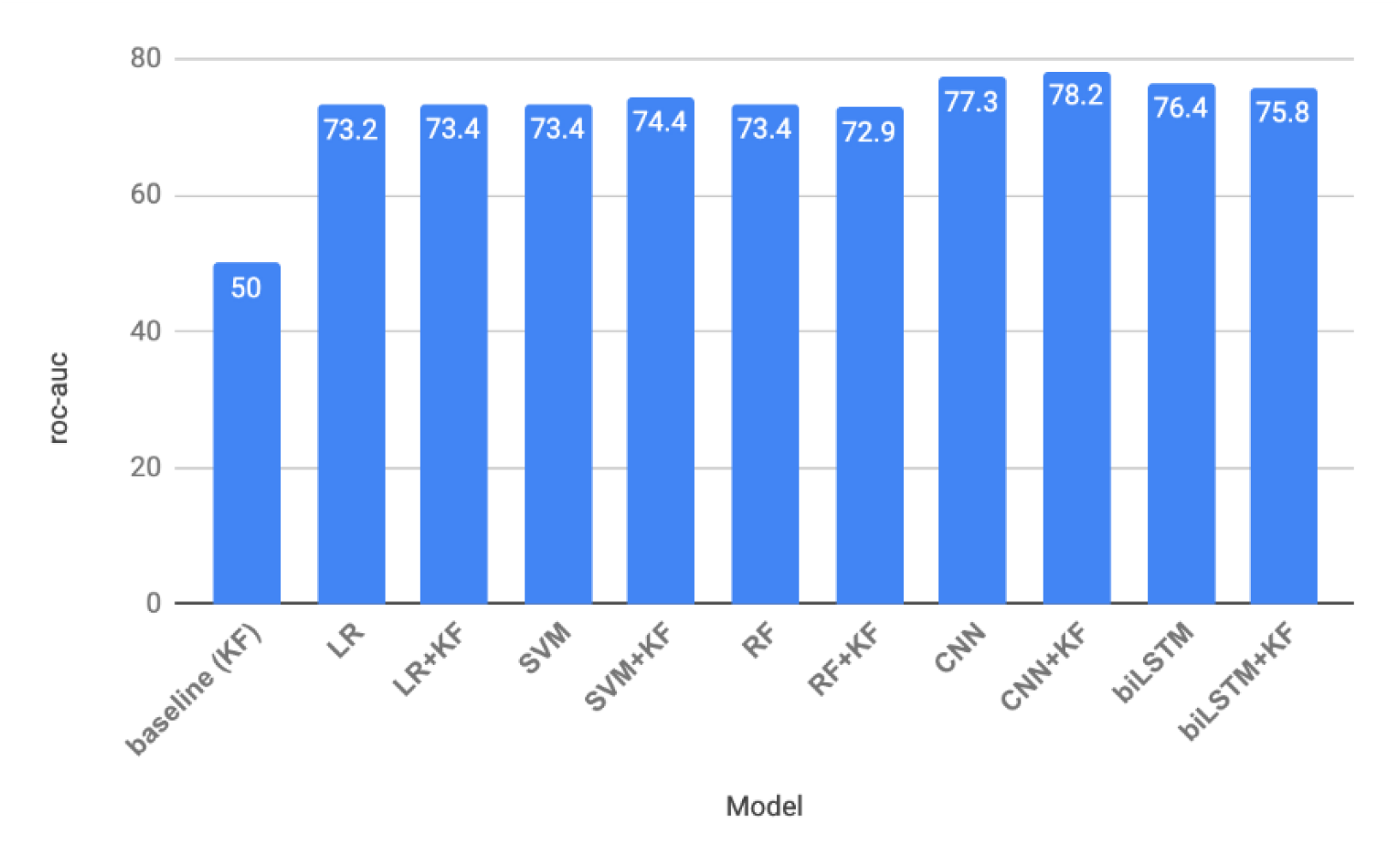
AUC-ROC vs. ML models for classifying irAEs.

Performance of the classifiers for predicting the top three irAEs subtypes are typically lower but comparative performance of models with and without filtering is similar to results obtained for classifying any io-toxicity i.e. deep learning models with word embeddings and keyword filtering outperform other models. For classifying skin-related toxicities, CNN with KF yielded the best F1-score of 65.1% (Table 3a). biLSTM with keyword filtering and class weight parameter was the best model for detecting endocrine toxicity with an F1-score of 58.8% (Table 3b). For colitis, the best results were obtained by CNN with keyword filtering and class-weights with an Ff1-score of 70.9% (Table 3c).

### Scattertext Results

Scattertext analysis (Fig 4) outlines the top words present in clinical notes with irAE and not irAE. The tool analyzed around three thousand words from the text corpus to assign a scaled f-score to each word. Ranking words by f-score can help identify which terms are more characteristic of irAE versus not irAE. Top words used in clinical notes of patients of having developed an irAE were: “prednisone”, “hydrocortisone”, “endocrinology”, “taper”, “steroids”, “hypothyroidism”, “nivo”, “cream”, “ipilimumab”, “steroid”, “benadryl”, “pruritic”, “pruritus”, “immune”. Top words used in clinical notes of patients of not having developed any irAE were: “dilaudid”, “oxycodone”, “chemotherapy”, “hemoptysis”, “oxycontin”, “radiation”, “progressive”, “exertion”, “productive”, “palliative”, “regimen”, “patch”, “fentanyl”.

**Fig 4.**
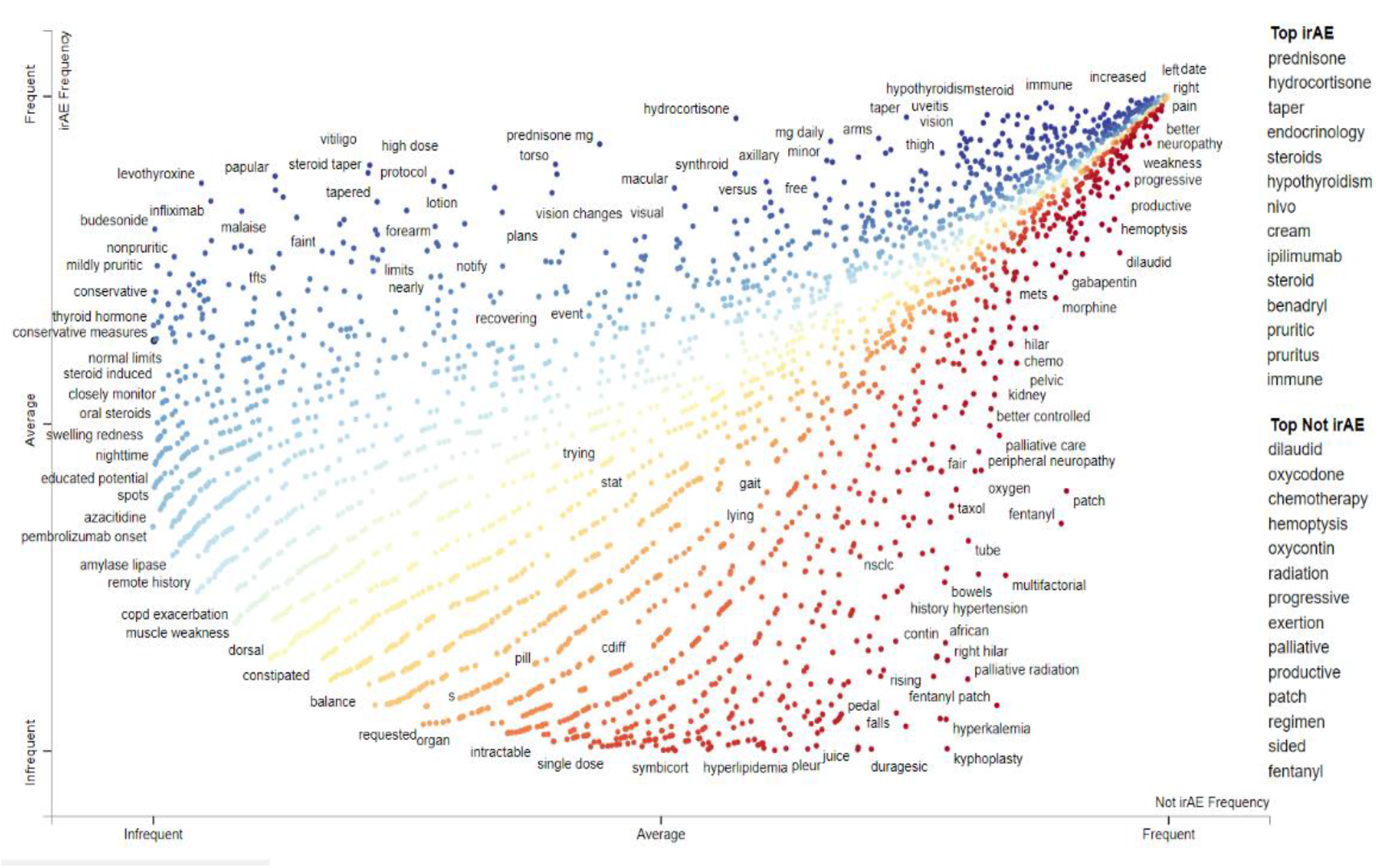
Scattertext visualization of words associated with irAE and no irAE groups.

The red dots on the plot represent terms that are more associated with irAEs compared to the blue dots which indicate terms more associated with no irAEs.

## Discussion

We analyzed some of the false positives (FPs) and false negatives (FNs) made by the deep learning models when classifying patient’s clinical notes for irAEs. Majority of FPs involve cases where the text for a patient contains several mentions of adverse events due to some other reason and not due to the ICI therapy. A majority of FNs involve cases where multiple toxicities might be mentioned in the set of notes for a positive patient, but only one toxicity is asserted, while the others mentioned in negative, speculative, resolved or cautionary context. Example 4, depicts one such case, which shows text snippets from the clinical notes of a patient with the irAE “pneumonitis”.

### Example 4

… *she appears to be tolerating well other than faint rash, some fatigue and dry mouth .….she has a* ***mild (gr1) rash*** *at this time but this appears to be* ***her only toxicity*** *.….we reviewed the potential side effects of pembrolizumab, including but not limited to, fatigue, pneumonitis, endocrinopathies, hepatitis, colitis, and rash …..sore throat persists but she has no s/s infection on physical exam*.

In the above example, there are several sentences with irAEs mentioned, but only one sentence relevant for classification. Since we are combining all sentences from all clinical notes, such “noisy” sentences dominate the combined text for a patient and a one layer-CNN architecture with the entire text in the input layer might not be able to discriminate such cases effectively. A possible solution might be to further filter text by removing sentences with negated, hedged, resolved or cautionary mentions of irAEs. Further hierarchical deep learning architecture with word and sentence attention layers [38] could help with such FN cases by enabling it to attend differentially to important content when constructing the text representation.

A large proportion of patient information such as adverse events to drugs/treatments, patient-reported outcomes, family history exists in unstructured text format in clinical notes. For example, adverse events to ICIs are buried in unstructured clinical notes and very infrequently recorded in a patient’s structured EHR. A review of the MedStar EHR system for our cohort indicated that only 16.7 % of patients who developed an irAEs had an adverse event (not necessarily due to ICI) recorded in their EHR. A manual chart review for annotating evidence of a patient phenotypes such as irAEs is costly and time-consuming. Automatically extracting such patient information from the clinical text can lead to a better understanding of health and diseases.

Such automatic deep phenotyping of a patient based on the clinical text will be helpful to identify patients who have developed the medical condition (phenotype) in question. Additionally, such NLP systems help create annotated datasets by reducing the clinician’s effort for manual chart review. These curated datasets could be used to develop predictive models to identify patients who are at risk of developing adverse events or predict a patient’s trajectory following treatment. Such automated deep phenotyping systems could also be useful in identifying patients for clinical trials, conducting pharmaco safety studies and comparing tests and treatments. In this work, we demonstrated that NLP methods based on deep learning models can achieve good performance for detecting immunotherapy toxicities from clinicians’ notes. In the future, we plan to extend this work to detect toxicity grades and immunotherapy outcomes (e.g. “no response”, “partial response”, “full response”), thereby building a comprehensive deep phenotyping detection system. This, in conjunction with structured data in the EHR, will enable extraction of the patient’s complete trajectory following treatment.

## Conclusion

In the real world setting, the evidence of irAEs are buried in clinical narratives and rarely captured in the structured format. portion of a patient’s EHR. In this work, we present a machine learning approach to classify patients treated with immunotherapy as developing adverse events (irAEs) or not based on their longitudinal set of clinical notes. We compare the performance of different ML models (shallow and deep learning models) with different feature representations (frequency-based, distributed word embeddings) for patient-level irAE classification tasks. Our results indicate that deep learning models, particularly convolutional neural networks (F1-score: 75.2%) with pre-trained word embeddings outperform a naive rule-based (keyword search) approach as well as shallow ML models for detecting any irAE. Additionally, we also evaluated the performance of ML models in detecting the top three irAEs in our dataset: skin-related toxicities, endocrine toxicities, and colitis. Results indicate that deep learning models perform best while classifying the top three irAE subtypes, with F1-scores of 65.1%, 58.8% and 70.9 % for skin-related toxicity, endocrine toxicity, and colitis respectively.

## Data Availability

The parent study was conducted under an IRB study #2017-0559 approved by
Georgetown University review board. The current project used a de-identified dataset
from the registry and didn't require an IRB.

